# Steep increases in fentanyl-related mortality west of the Mississippi River: synthesizing recent evidence from county and state surveillance

**DOI:** 10.1101/2020.06.15.20131839

**Authors:** Chelsea L. Shover, Titilola O. Falasinnu, Candice L. Dwyer, Nayelie Benitez Santos, Nicole J. Cunningham, Noel A. Vest, Keith Humphreys

## Abstract

**Background and Aims:** Overdose deaths from synthetic opioids (e.g., fentanyl), increased 10-fold in the United States from 2013-2018, despite 88% of deaths occurring east of the Mississippi River. Public health professionals have long feared that further spread of fentanyl could greatly exacerbate the opioid epidemic. We aimed to measure and characterize recent fentanyl deaths in jurisdictions west of the Mississippi River.

**Design:** Systematic search of states and counties in the Western U.S. for publicly available data on fentanyl-related deaths since the most recently published Centers for Disease Control and Prevention (CDC) statistics, which cover through December 2018. Longitudinal study using 2019 and 2020 mortality records to identify changes in fentanyl-involved mortality since most recent CDC statistics.

**Settings:** U.S. states west of the Mississippi River.

**Measurements:** Annual rate of fentanyl-involved deaths per 100,000 population. Proportion of fatal heroin-, stimulant, and prescription pill overdoses also involving fentanyl.

**Findings:** We identified nine jurisdictions with publicly available fentanyl death data through December 2019 or later - State of Arizona; Denver County, CO; Harris County, TX; Humboldt County, CA; King County, WA; Los Angeles County, CA; San Francisco County, CA; Siskiyou County, CA; Dallas-Fort Worth, TX metro area (Denton, Johnson, Parker, Tarrant counties. Fentanyl deaths increased in each jurisdiction. Their collective contribution to national synthetic narcotics mortality tripled from 2017 to 2019. First quarter 2020 data (available from all but San Francisco County) showed a 33% growth in fentanyl-mortality over 2019. Fentanyl-involvement in heroin, stimulant, and prescription pill deaths has grown substantially over time.

**Conclusions:** Fentanyl has spread westward, which could dramatically worsen the nation’s already severe opioid epidemic. Increasing standard-dose of naloxone, expanding Medicaid, improving coverage of addiction treatment, and public health educational campaigns should be prioritized.

## INTRODUCTION

Since 2013, deaths involving synthetic opioids – mainly fentanyl and its analogues – have increased ten-fold in the United States, with over 31,000 deaths nationwide in 2018.(1-5) Remarkably, this carnage has showed up in national statistics despite fentanyl-penetration of illicit drug markets being largely confined to the Eastern U.S.(6, 7) In 2018, the 28 states east of the Mississippi River accounted for 88% of synthetic opioid overdose deaths.(8) As recently as summer 2019, drug seizure and mortality data suggested that illicitly manufactured fentanyl remained almost entirely concentrated east of the Mississippi River, raising hopes that this deadly drug would not gain a national foothold.(6, 7, 9) The spread of deadly drugs across illicit markets is by no means inevitable. Estonia for example has had a fentanyl-dominated illicit opioid market for two decades whereas Finland, just a short ferry ride away, does not.(7, 10) Sometimes cultural norms, market dynamics, and law enforcement manage to constrain a particularly deadly drug to one region. If this happened in the U.S., it would lessen the national death toll.

However, research published in 2020 indicates that fentanyl has become nearly ubiquitous in heroin samples (as well as to a lesser extent in cocaine and methamphetamine) evaluated just across U.S. borders both in Northwestern Mexico(11) and Western Canada.(12) News reports of increasing fentanyl overdoses in late 2019 and early 2020 in various U.S. jurisdictions west of the Mississippi River raise further concern,(13-18) as do reports indicating that fentanyl supply has been largely unimpacted by the novel coronavirus pandemic.(19, 20) Given how fentanyl has so dramatically worsened the U.S. overdose death rate while only being pervasive in part of the country, its national spread could make the epidemic significantly worse.

The Centers for Disease Control and Prevention (CDC)’s most current provisional national synthetic opioid overdose mortality statistics cover October 2019,(21) yet the most currently available state-level statistics only reach through December 2018.(1) The substantial lag in availability of state-level mortality data makes surveillance of regional emerging drugs trends difficult and thereby reduces the ability of public health officials to respond rapidly.

To investigate the degree to which fentanyl has recently penetrated drug markets west of the Mississippi River, we synthesized mortality data from local and state health departments and medical examiner offices. We primarily report changes in population-level rate of fentanyl mortality in the time since the most recently available CDC data. We additionally utilized this data to investigate changes in proportion of heroin-, methamphetamine-, cocaine-, and pill- (prescription opioids, benzodiazepines, 3,4-methylenedioxymethamphetamine) involved deaths that also involve fentanyl.

## METHODS

We use recent data from national, state, and county sources to investigate potential changes in fentanyl-involved deaths west of the Mississippi. The primary outcome was changes in fentanyl-involved deaths per 100,000 population in each jurisdiction. The secondary outcome was proportion of deaths involving other drugs (methamphetamine, cocaine, heroin, prescription opioids, benzodiazepines) that also involved fentanyl. Owing to the fragmented nature of immediately available state and local health jurisdiction data, the data collection strategy was as systematic as possible to collect what are ultimately unsystematic results. The tradeoff of results obtained this way is that variation in time frame and level of detail is compensated for by greater immediacy and specificity relative to national data currently available from the CDC.(1, 22)

### Data sources

Data were included from the 22 states that are entirely west of the Mississippi River (Alaska, Arkansas, Arizona, California, Colorado, Hawaii, Idaho, Iowa, Kansas, Missouri, Montana, North Dakota, Nebraska, New Mexico, Nevada, Oklahoma, Oregon, South Dakota, Texas, Utah, Washington, Wyoming). For each state, we searched the state and county health department websites for 1) overdose surveillance data that reported fentanyl-involved deaths 2) public medical examiner data. We identified states and counties that reported fentanyl-involved mortality from 2019 or later (**Figure 1**).

**Figure 1.**
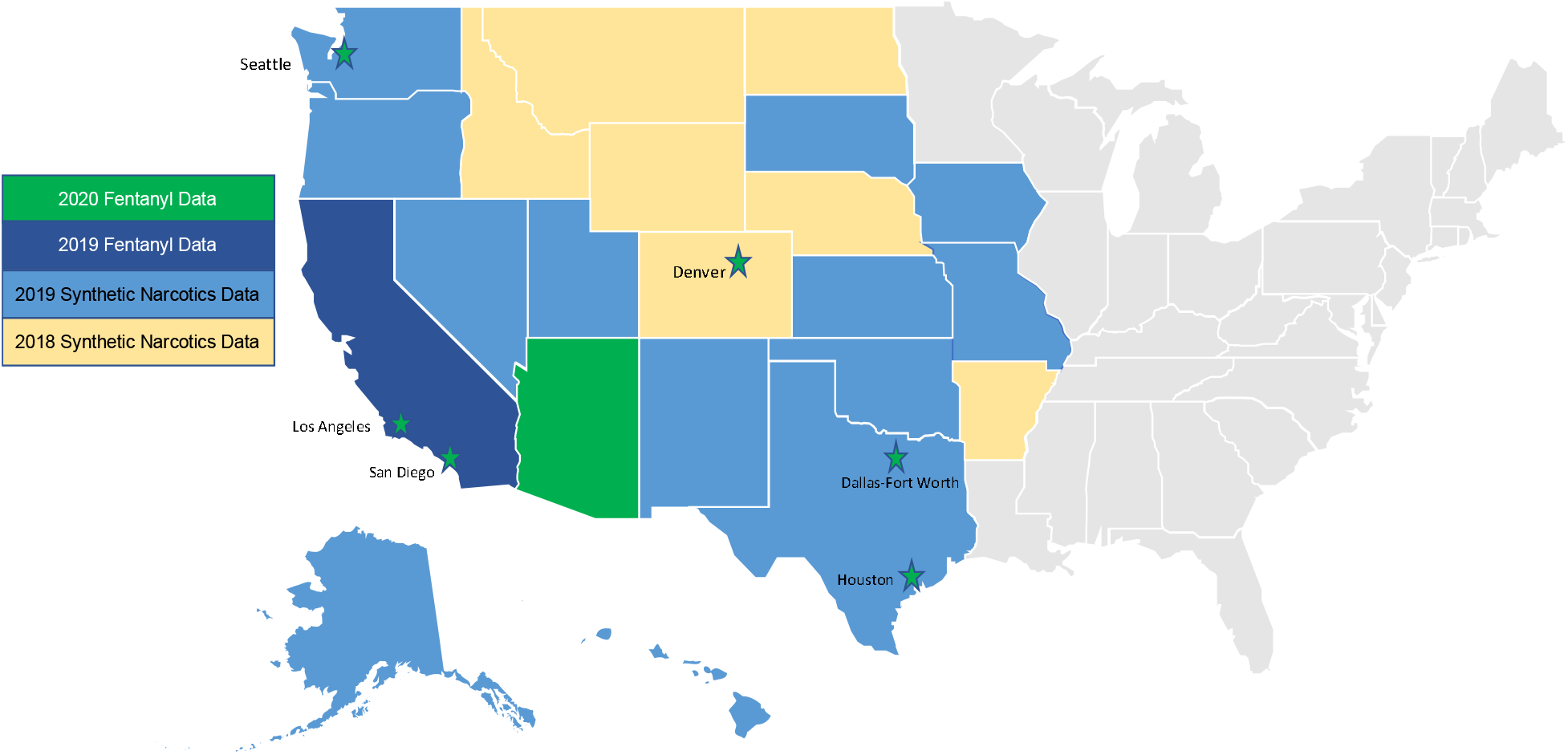
Most recently available fentanyl or synthetic opioid mortality data for jurisdictions west of the Mississippi River.

We searched each state’s health department website for a state-level opioid dashboard. Two states – Arizona and California – report fentanyl-involved overdoses through state dashboards, with Arizona reporting fatal fentanyl overdoses until the current month (April 2020) and California reporting fatal fentanyl overdoses through June 2019.(23, 24) Local health department websites were identified from the National Association of County Health Officials directory and cross-checked with the state’s public health department website. For each state, we also searched for jurisdictions with publicly available online medical examiner case data.

In addition to the two states, our review of state and local health departments identified 11 counties and one metro area that provided a public report of count of fentanyl-involved deaths in 2019 or 2020 or individual-level medical examiner data necessary to create such reports.(13, 18, 25-29) Individual death records were available from Los Angeles County, CA;(30) San Diego County, CA;(31) Denver County, Co;(32) Harris County, TX (Houston metro area);(33) Dallas-Fort Worth, TX (comprised of four counties);(34) Maricopa County, AZ.(35)

San Diego and Dallas-Fort Worth had publicly available exportable death records over multiple years, which allowed us to efficiently measure changes in fentanyl-involved deaths over time. Denver County provided an aggregate file for 2019’s drug-related deaths. For jurisdictions with publicly available individual death records (Los Angeles, Harris County), we manually reviewed all medical examiner case records from December 2019 through March 2020, and the earliest available month from 2019 as a comparison for the drug combination analysis. We reviewed 2,858 individual records from Los Angeles County and 1,887 in Harris County. Because Maricopa County is located within a state that provides statewide monthly counts of fentanyl overdoses, we reviewed only one month (January 2020, 477 individual records) in order to capture a snapshot of drug combinations.

To investigate change in these jurisdictions’ contribution to national synthetic opioid overdose mortality, we extracted multiple cause of death data from 2018 and earlier through the CDC’s Wide-ranging Online Data for Epidemiologic Research multiple cause mortality file.(1) We extracted deaths that included *International Classification of Diseases 10*^*th*^ *Edition* Code T40.4, “other synthetic narcotics,” a category that includes fentanyl and its analogues along with tramadol, meperidine and tapentadol, among others but excludes methadone.(36) Provisional national data through October 2019 was obtained through the CDC’s Vital Statistics Rapid Release online portal.(21)

### Statistical Methods

To compare across cities and states with widely varying populations, annual rate per 100,000 population was calculated based on the number of fentanyl-involved deaths and population estimates from U.S. Census Bureau Quick Facts.(37) When an entire year’s data were not available, the annual rate was imputed based on the average of the year’s available months. To contextualize findings within the national opioid overdose epidemic, we calculated the proportion of synthetic opioid overdose deaths that the reviewed jurisdictions contributed to the national total in 2017 and 2018 using CDC mortality data. We estimated the 2019 contribution using the national total from the year-ending October 2019. The research questions were not pre-registered and results should therefore be considered exploratory.

### Ethics

This study was designated as exempt from Institutional Review Board oversight by the [blinded] IRB.

## RESULTS

The review of states and counties identified 14 jurisdictions with any fentanyl death data more recent than the CDC data. Increases in fentanyl-involved mortality since the CDC’s most recently released data were observed in all 14 jurisdictions (**Table 1**).

**Table 1.** Number of annual fentanyl-involved deaths by jurisdiction with publicly available data more current that most recently released Centers for Disease Control and Prevention data

| Jurisdiction | Most recent data | 2019 Population | 2017 |  | 2018 |  | 2019 |  | 2020 (Projected) |  | Annual change |  |  |
| --- | --- | --- | --- | --- | --- | --- | --- | --- | --- | --- | --- | --- | --- |
|  |  |  | n | Rate/100,000 pop | n | Rate/100,000 pop. | n | Rate/100,000 pop. | n | Rate/100,000 pop. | 2017-18 | 2018-19 | 2019-20 (Project) |
| Arizona (statewide) | Apr. 2020 | 7,278,717 | 239 | 3.4 (3.0, 6.4) | 648 | 9.0 (8.3, 17.4) | 1,192 | 16.4 (15.4, 31.8) | 1,374 | 18.9 (17.9, 36.8) | 171% | 84% | 13% |
| California (statewide) | Jun. 2019 | 39,512,223 | 431 | 1.1 (1.0, 2.1) | 786 | 2.0 (1.8, 3.8) | 1,007 | 2.5 (2.4, 4.9) | -- | -- | 82% | 28% | -- |
| Dallas/Fort-Worth, TX | Apr. 2020 | 3,308,417 | 19 | 0.6 | 10 | 0.3* | 16 | 0.5* | 24 | 0.7 (0.4, 1.2) | -47% | 60% | 50% |
| Denver County, CO | Mar. 2020 | 727,211 | 18 | 1.4* | 17 | 2.4* (1.2, 3.6) | 53 | 7.3 (5.3, 12.6) | 45 | 6.2 (4.4, 10.6) | -6% | 212% | -15% |
| Harris County, TX | Mar. 2020 | 4,713,325 | 59 | 1.3 (0.9, 2.2) | 97 | 2.1 (1.7, 3.7) | 102 | 2.2 (1.7, 3.9) | 172 | 3.6 (3.1, 6.8) | 64% | 5% | 69% |
| Humboldt County, CA | Dec. 2019 | 135,558 | 1 | 0.7* | 0 | 0.0 | 7 | 5.2* | -- | -- | -- | >600% | -- |
| King County, WA | Apr. 2020 | 2,252,782 | 33 | 1.5 (1.0, 2.4) | 66 | 2.9 (2.2, 5.2) | 112 | 6.0 (4.1, 9.0) | 152 | 6.7 (6.2, 13.0) | 100% | 70% | 36% |
| Los Angeles County, CA | Mar. 2020 | 10,039,107 | 117 | 1.2 (0.9, 2.1) | 201 | 2.0 (1.7, 3.7) | 378 | 3.8 (3.4, 7.2) | 676 | 6.7 (6.2, 13.0) | 72% | 88% | 79% |
| Maricopa County, AZ | Apr. 2020 | 4,485,414 | -- | -- | -- | -- | -- | -- | 900 | 20.1 (18.8, 38.8) | -- | -- | -- |
| San Diego County, CA | Jun. 2019 | 3,338,330 | 73 | 2.2 (1.7, 3.9) | 87 | 2.6 (2.1, 4.7) | 130 | 3.9 (3.2, 7.1) | -- | -- | 19% | 49% | -- |
| San Francisco County, CA | Dec. 2019 | 881,549 | 36 | 4.1 (2.7, 6.8) | 89 | 10.1 (8.1, 18.1) | 162 | 18.4 (15.5, 33.9) | -- | -- | 147% | 82% | -- |
| San Luis Obispo County, CA | Oct. 2019 | 283,111 | 0 | 0 | 1 | 0.4* | 16 | 5.7* | -- | -- | -- | 1500% | -- |
| Santa Cruz County, CA | Aug. 2019 | 273,213 | 1 | 0.4* | 2 | 0.7* | 17 | 5.5* | -- | -- | 100% | 750% | -- |
| Siskiyou County, CA | Dec. 2019 | 43,539 | 1 | 2.3* | 0 | 0.0 (0.0, 0.0) | 1 | 2.3* | -- | -- | -100% | >100% | -- |

\*Rate is calculated based on fewer than 20 cases and is thus unreliable

Of these, data through December 2019 or later was available for one state (Arizona), one metro area (Dallas-Fort Worth, TX, including Tarrant, Denton, Parker, and Johnson Counties), and seven counties: Denver County, CO; Harris County, TX; Humboldt County, CA; King County, WA; Los Angeles County, CA; San Francisco County, CA; Siskiyou County, CA; These nine jurisdictions comprise 9% of the total United States population in 2019. In 2017, fentanyl deaths across these nine jurisdictions (522) contributed 1.8% of the national synthetic opioid overdose mortality (28,453). In 2018, the nine jurisdictions had 1,128 fentanyl deaths, which was 3.6% of the national count (31,327). In 2019, these jurisdictions had an estimated 2,021 fentanyl deaths, which was equivalent to 6.0% of the national synthetic opioid overdose mortality in the 12-months period ending in October 2019 (34,192). Among the six jurisdictions with data from 2019 and 2020, the fentanyl-involved mortality rate had increased 33% in 2020 over 2019, with steepest increases in the Los Angeles (82%) and Harris (73%) Counties.

Among 14 jurisdictions west of the Mississippi River, the highest annual fentanyl-related deaths per 100,000 population was observed in Maricopa County, AZ in January 2020 (20.1, 95% CI 18.8, 36.8); followed by Arizona statewide in January-April 2020 (18.9 (17.9, 36.8); San Francisco County, CA in 2019 (18.4, 95% CI, 15.5, 33.9); Denver County, CO in 2019 (7.3, 95% CI 5.3, 12.6); King County, WA in January-March 2020 (6.7, 95% CI 5.7, 12.4); and Los Angeles County, CA in January through March 2020 (6.9, 95% CI 6.3, 13.2) (**Table 1, Figure 1**).

Other California jurisdictions also had notable increases in fentanyl death-rate, with the largest in Santa Cruz, San Luis Obispo, Humboldt, and San Diego counties. In Texas, rates varied considerably between major cities. Harris County had a fentanyl death rate of 3.7 (95% CI, 3.2, 6.9) per 100,000 in early 2020, while the four counties of the Dallas-Fort Worth area consistently had a fentanyl-involved death rate of only 0.6 (95% CI, 0.4, 1.0) per 100,000 or less through April 2020.

San Diego County medical examiner data provided the longest time horizon to examine fentanyl involvement in deaths involving other substances. Proportion of heroin-involved deaths in San Diego County with fentanyl involvement grew from 0% in 2014 to 20% in the first half of 2019 (**Figure 2**). Similarly, while in 2014 no cocaine deaths involved fentanyl, in 2018 and the first half of 2019 fentanyl was involved in 39% and 33% of cocaine-related deaths respectively.

**Figure 2.**
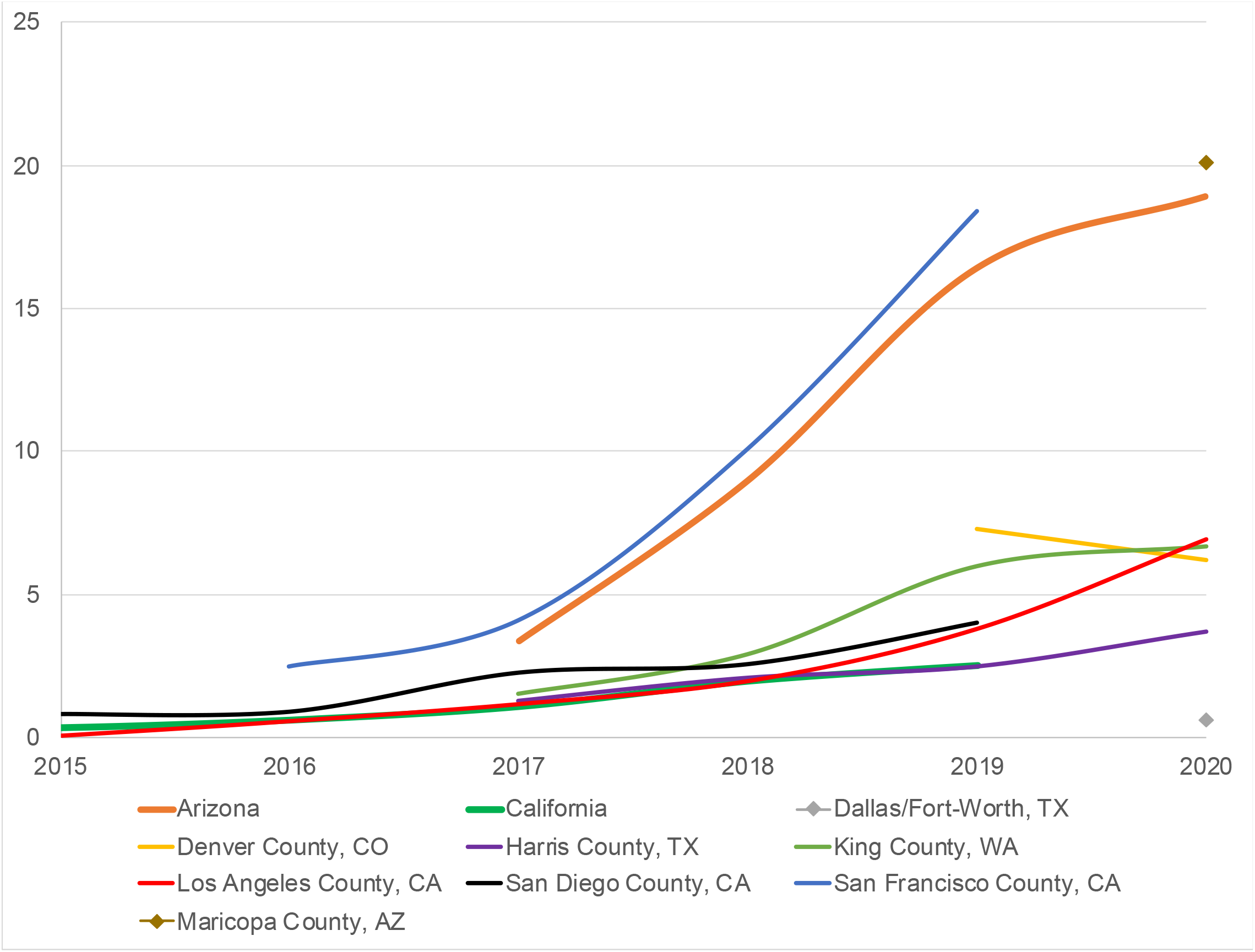
Annual rate of fentanyl-related deaths per 100,000 population.

Los Angeles County’s 2017 comparison data from the National Drug Early Warning System differ in including toxicology results for all deaths, including those where a substance was present but not a cause of death, whereas the 2019 and 2020 data we collected includes only drugs listed as a cause of death.(38) That said, the same pattern was evident, as from 2017 to 2020, the prevalence of fentanyl grew from 15% to over 60% in cocaine cases and from 6% to over 40% in methamphetamine cases.

In Harris County, the proportion of other drug-related deaths involving fentanyl varied over time, whereas in Dallas-Fort Worth the proportions increased but remained lower than the other cities overall, with the percentage of heroin-related deaths also involving fentanyl never surpassing 6%.

In January 2020, fentanyl was involved in 44% (n=75) of drug-related deaths in Maricopa, AZ, and in 62% (n=10) cocaine-related deaths, 31% (n=19) of methamphetamine-related deaths, and 25% (n=10) of heroin-related deaths. In 2019 in Denver, CO, 23% (n=53) of all drug-related deaths involved fentanyl, as did 12% of both methamphetamine (n=9) and cocaine-related (n=5) deaths. In 2019, only 6% (n=6) of heroin-related deaths in Denver involved fentanyl.

The review of county health department websites identified reports of counterfeit pressed pills containing fentanyl, such as oxycodone or alprazolam(14, 17, 39-42) as well as reports of heroin laced with fentanyl.(16, 43) Although alprazolam was involved in a relatively low number of overdoses, the proportion with fentanyl-involvement was typically high. Across all cities with data available for 2020, 30 (49%) of 61 alprazolam deaths also involved fentanyl.

We constructed a Venn diagram of drug categories to illustrate fentanyl’s involvement in other fatal overdoses in markets with substantial fentanyl penetration and 2020 data (Maricopa County, AZ; Los Angeles County, CA; and Harris County, TX) (**Figure 3**). There were 631 fatal overdoses that involved one of more of the following categories: fentanyl (fentanyl or fentanyl analogs), heroin (heroin or morphine), pills (semi-synthetic opioids –i.e., oxycodone, hydrocodone, hydromorphone, oxymorphone – benzodiazepines, or 3,4-methylenedioxymethamphetamine), or stimulants (methamphetamine, cocaine, amphetamines). Overlap between fentanyl and each of these categories was substantial.

**Figure 3.**
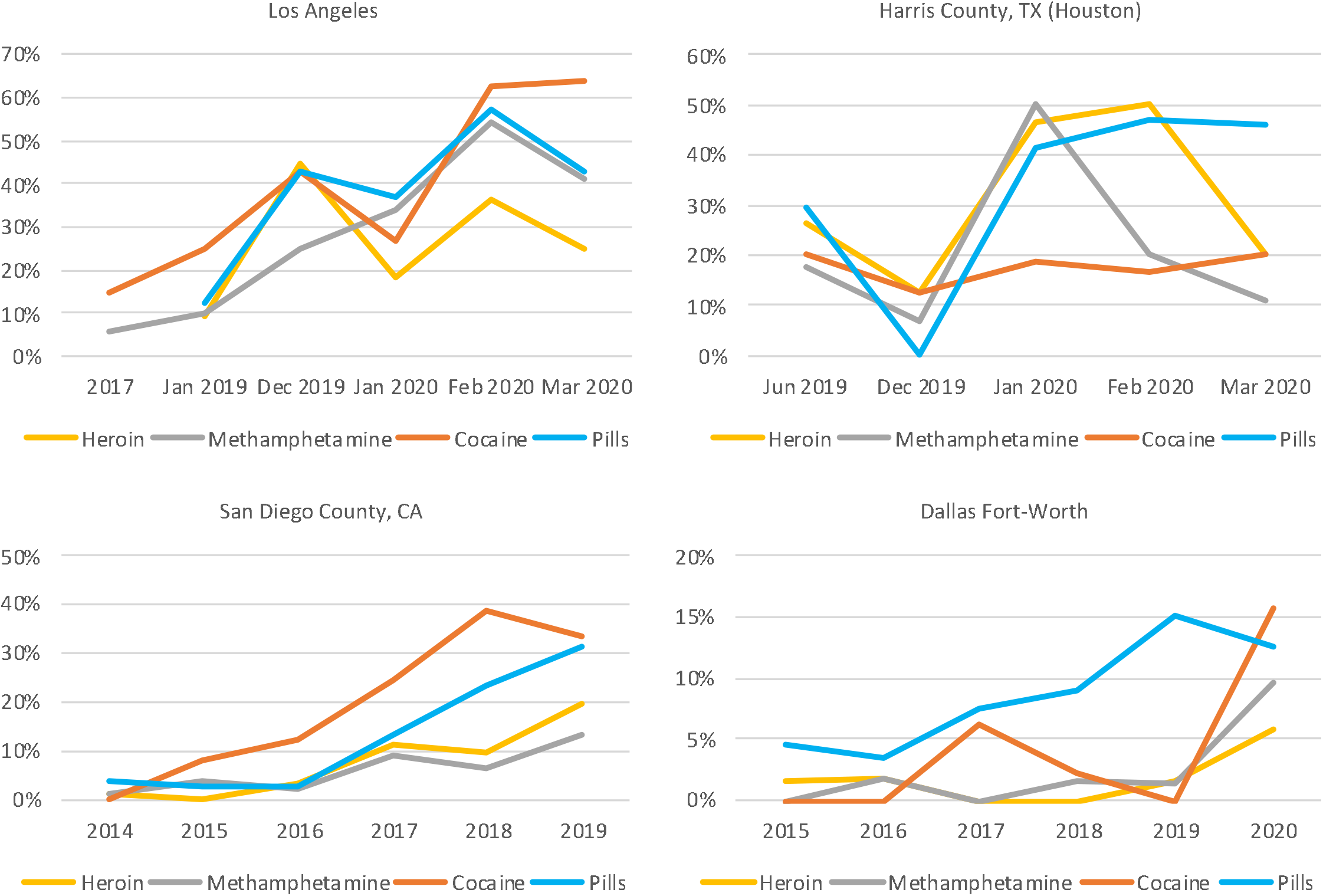
Proportion of drug-involved deaths also fentanyl positive. Legend: Fentanyl includes fentanyl and fentanyl analogs. Heroin includes heroin and morphine. Pills include semi-synthetic opioids –i.e., oxycodone, hydrocodone, hydromorphone, oxymorphone – benzodiazepines,3,4-methylenedioxy-methamphetamine (MDMA)).

**Figure 4.**
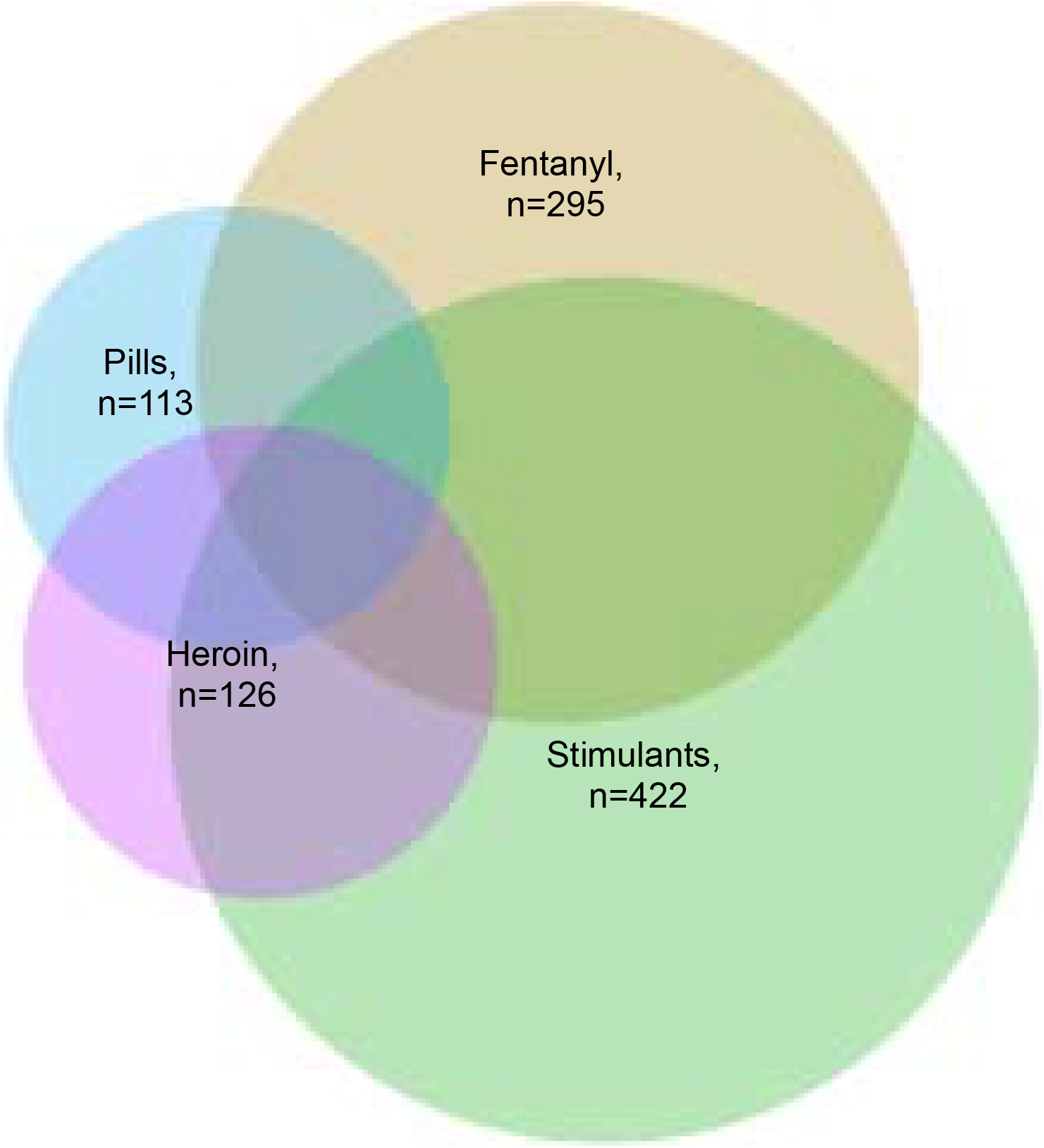
Drug combinations in fatal overdoses in three jurisdictions west of the Mississippi River with substantial fentanyl penetration in 2020, n=631. Legend: Data from Harris County (Jan-Mar 2020), Los Angeles County (Jan-Mar 2020), Maricopa County (Jan 2020). Fatal overdoses that involve one or more of the following are included: Fentanyl (fentanyl or fentanyl analogs), Heroin (heroin or morphine), Pills (semi-synthetic opioids –i.e., oxycodone, hydrocodone, hydromorphone, oxymorphone – benzodiazepines,3,4-methylenedioxy-methamphetamine (MDMA)), or Stimulants (methamphetamine, cocaine, amphetamines).

## DISCUSSION

We report a marked increase in the rate of fentanyl-related deaths west of the Mississippi River in every jurisdiction where such data were available. Moreover, we observed increases in the proportion of both heroin and stimulant (methamphetamine and cocaine) fatal overdoses that also involved fentanyl. This suggests increasing, worrisome, penetration of these markets by fentanyl. These changes are not yet reflected in CDC data and hence have not, to our knowledge, come to policymakers’ attention. Enhancing awareness of fentanyl’s deadly spread is urgent, because it can stimulate a rapid response before this growing public health crisis worsens.

The share of U.S. synthetic opioid overdose mortality attributable to the Western jurisdictions from which we gathered data more than tripled from 2017 to 2019, even as the overall synthetic opioid overdose mortality grew. Furthermore, increases from 2019 into early 2020 in Arizona, Los Angeles, Seattle, and Houston suggest that growth may continue.

2018 saw the first drop in overall drug overdose mortality in 40 years, but the spread of fentanyl is likely to more that reverse that – a speculation that CDC’s provisional overdose data support. The year-ending October 2019 shows 47,852 opioid-involved deaths nationally, up from 47,273 in the year-ending October 2018.(21) Of these, 34,192 involve synthetic opioids, an absolute increase of 2,930 deaths (9%) over the previous year. Continuing to monitor the CDC data is important, but we cannot afford to wait until that data are available to take action necessary to prevent more deaths.

The results have several important policy and practice implications. In markets where fentanyl has fully penetrated the heroin supply, distributing fentanyl testing strips to heroin users is now as useful as advising testing of drinking water for the presence of hydrogen. Instead, the most accurate public health message would be to communicate that to use a drug described as heroin is to almost certainly use fentanyl. In contrast, because of dealer and user market differentiation as well as high-level illicit production dynamics, fentanyl testing strips may still be useful for cocaine and methamphetamine users as the penetration of fentanyl in those drugs may never be total. Similarly, test strips may be useful for pills obtained without a prescription, as fentanyl has been widely but not universally observed in counterfeit pressed pills sold to resemble oxycodone, alprazolam, and other prescription medications.

The increasing prevalence of fentanyl in the form of lookalike pharmaceutical pills appears to be contributing to the deaths observed in the data.(17, 26, 27, 42, 44) The unfortunate history of scare-mongering in drug prevention messages (e.g., in mass media) now co-exists with a present reality in which “one pill can kill” warnings could be accurate. Public health departments, media, and schools, have a responsibility to broadcast this change in a fashion that neither overstates nor understates the risks posed.

Most of the recommendations to deal with fentanyl have already been made to deal with the prescription opioid and heroin addiction.(45, 46) But all of them now have an added urgency, particularly ensuring that all public and private insurers cover treatment, and that all FDA-approved medications for opioid use disorder are available in specialty and non-specialty health care settings, including correctional facilities. This would also be a propitious time to expand U.S. research on medication options for opioid use disorder with evidence of effectiveness in other countries, such as hydromorphone, slow-release oral morphine, and injectable methadone. Increasing the standard dose of naloxone is also warranted because overdoses involving synthetic opioids like fentanyl and fentanyl analogues often require more naloxone to reverse.(47, 48)

The stark differences between immediate and older surveillance data point to the need to implement innovative methods to track emerging drug trends. Rapid surveillance using wastewater is one option.(49) As we demonstrate, medical examiner case reports can be a rich source of data, but few are readily available and aggregating them is time-intensive.

### Limitations

Jurisdictional differences in reporting – including time period, outcome (fatal only vs. all fentanyl overdoses), drug category (synthetic opioids generally vs. fentanyl specifically) – make comparisons challenging. Changes within jurisdiction of data availability may add imprecision to results. For example, Los Angeles had 2017 drug combination data for all medical examiner cases with positive toxicology results, even if the cause of death was not drug-related. However, the individual records that comprise the 2019 and 2020 Los Angeles data only contain information on drugs implicated in death.

Using mortality data inherently underestimates the total number of fentanyl overdoses, since non-fatal overdoses are not systematically captured in any jurisdiction. Mortality data could simultaneously overestimate the degree of penetration of fentanyl into the supply of other drugs, if stimulants or heroin that contain fentanyl are more likely to result in a fatal overdose compared to the same drugs without fentanyl. Due to variation in medical examiner investigation time, some records were pending as of this writing: 93 (3%) in Los Angeles County, and 61 (3%) in Harris County. As these cases are resolved, the number of deaths attributed to fentanyl will likely increase.

Our projections for 2020 where available are simplified; they show what the year-end death rate would be if the rate observed in the available 2020 data continues for the rest of the year. With a few exceptions, the rate of fentanyl deaths in each jurisdiction has grown over time; therefore, this strategy likely underestimates the true death toll. Similarly, in some cases – Los Angeles, Harris County, San Diego County, Santa Cruz County, San Luis Obispo County, the State of California, the values reported for 2019 are extrapolated from the available months. In jurisdictions where the only available data were from January through June (California, San Diego), this may underestimate the annual total if deaths increased over the year. Conversely, it is possible that Harris County (2019 total is six times sum of June and December deaths) and Los Angeles County (2019 total is January through June total plus six times December deaths) overestimates the annual total by more heavily weighting later months in the year. However, the counts observed in January through March 2020 show substantial increases that suggest such overestimation is unlikely.

### Conclusion

As evidenced by mortality data from as recently as the first quarter of 2020, fentanyl is not only meaningfully present west of the Mississippi River, but appears to be increasing dramatically in nearly all jurisdictions with timely data. If public health officials do not react strongly and quickly, a significant exacerbation of the U.S. opioid epidemic could follow.

## Data Availability

Data will be available to peer reviewers and available on request after publication.

## ACKNOWLEDMENTS

CLS, TOF, and NV were supported by National Institute on Drug Abuse T32 DA035165. KH was supported by grants from the U.S. Veterans Health Administration. CLS and KH were supported by the Wu Tsai Neurosciences Institute. The funders had no role in design and conduct of the study; collection, management, analysis, and interpretation of the data; preparation, review, or approval of the manuscript; or decision to submit the manuscript for publication. The content is solely the responsibility of the authors and does not necessarily represent the official views of the NIH or the authors’ employers.

